# Clinical and immunologic features in severe and moderate forms of Coronavirus Disease 2019

**DOI:** 10.1101/2020.02.16.20023903

**Authors:** Guang Chen, Di Wu, Wei Guo, Yong Cao, Da Huang, Hongwu Wang, Tao Wang, Xiaoyun Zhang, Huilong Chen, Haijing Yu, Xiaoping Zhang, Minxia Zhang, Shiji Wu, Jianxin Song, Tao Chen, Meifang Han, Shusheng Li, Xiaoping Luo, Jianping Zhao, Qin Ning

**Affiliations:** Department and Institute of Infectious Disease, Tongji Hospital, Tongji Medical College, Huazhong University of Science and Technology, Wuhan, China; Department of Respiratory Disease, Tongji Hospital, Tongji Medical College, Huazhong University of Science and Technology, Wuhan, China; Department of Laboratory Medicine, Tongji Hospital, Tongji Medical College, Huazhong University of Science and Technology, Wuhan, China; Department of Emergency Medicine, Tongji Hospital, Huazhong University of Science and Technology, Wuhan, China; Department of Pediatrics, Tongji Hospital, Tongji Medical College, Huazhong University of Science and Technology, Wuhan, China

**Keywords:** SARS-CoV-2, COVID-19, cytokines, lymphocytes, IFN-γ

## Abstract

**Background:** Since late December, 2019, an outbreak of pneumonia cases caused by the severe acute respiratory syndrome coronavirus 2 (SARS-CoV-2) emerged in Wuhan, and continued to spread throughout China and across the globe. To date, few data on immunologic features of Coronavirus Disease 2019 (COVID-19) have been reported.

**Methods:** In this single-centre retrospective study, a total of 21 patients with pneumonia who were laboratory-confirmed to be infected with SARS-CoV-2 in Wuhan Tongji hospital were included from Dec 19, 2019 to Jan 27, 2020. The immunologic characteristics as well as their clinical, laboratory, radiological features were compared between 11 severe cases and 10 moderate cases.

**Results:** Of the 21 patients with COVID-19, only 4 (19%) had a history of exposure to the Huanan seafood market. 7 (33.3%) patients had underlying conditions. The average age of severe and moderate cases was 63.9 and 51.4 years, 10 (90.9%) severe cases and 7 (70.0%) moderate cases were male. Common clinical manifestations including fever (100%, 100%), cough (70%, 90%), fatigue (100%, 70%) and myalgia (50%, 30%) in severe cases and moderate cases. PaO2/FiO2 ratio was significantly lower in severe cases (122.9) than moderate cases (366.2). Lymphocyte counts were significantly lower in severe cases (0.7 × 10□/L) than moderate cases (1.1 × 10□/L). Alanine aminotransferase, lactate dehydrogenase levels, high-sensitivity C-reactive protein and ferritin were significantly higher in severe cases (41.4 U/L, 567.2 U/L, 135.2 mg/L and 1734.4 ug/L) than moderate cases (17.6 U/L, 234.4 U/L, 51.4 mg/L and 880.2 ug /L). IL-2R, TNF-α and IL-10 concentrations on admission were significantly higher in severe cases (1202.4 pg/mL, 10.9 pg/mL and 10.9 pg/mL) than moderate cases (441.7 pg/mL, 7.5 pg/mL and 6.6 pg/mL). Absolute number of total T lymphocytes, CD4^+^T cells and CD8^+^T cells decreased in nearly all the patients, and were significantly lower in severe cases (332.5, 185.6 and 124.3 × 10^6^/L) than moderate cases (676.5, 359.2 and 272.0 × 10^6^/L). The expressions of IFN-γ by CD4^+^T cells tended to be lower in severe cases (14.6%) than moderate cases (23.6%).

**Conclusion:** The SARS-CoV-2 infection may affect primarily T lymphocytes, particularly CD4^+^T cells, resulting in significant decrease in number as well as IFN-γ production, which may be associated with disease severity. Together with clinical characteristics, early immunologic indicators including diminished T lymphocytes and elevated cytokines may serve as potential markers for prognosis in COVID-19.

## Introduction

Coronaviruses (CoV) are a large family of respiratory viruses that can cause diseases ranging from the common cold to the Middle-East Respiratory Syndrome (MERS) and the Severe Acute Respiratory Syndrome (SARS)^1,2^, both of which are zoonotic in origin and induce fatal lower respiratory tract infection as well as extrapulmonary manifestations. The new coronavirus, designated as the severe acute respiratory syndrome coronavirus 2 (SARS-CoV-2), is a member of Beta-CoV lineage B, which was first identified in Wuhan, China by the Chinese Center for Disease Control and Prevention (CDC)^3,4^. Recent reports have provided evidence for person to person transmission of the SARS-CoV-2 in family and hospital settings ^5,6^. As of Feb 1, 2020, the number of SARS-CoV-2 cases globally had eclipsed 14000, exceeding the total number of SARS cases during the 2003 epidemic, and more than 300 people had now died. The outbreak of SARS-CoV-2-induced Coronavirus Disease 2019 (COVID-19) has put health authorities on high alert in China and across the globe.

It has been revealed that SARS-CoV-2 has a genome sequence 75% to 80% identical to the SARS-CoV and has more similarities to several bat coronaviruses^3^. Both clinical and epidemiological features of patients with COVID-19 have recently been reported, demonstrating that the SARS-CoV-2 infection can cause clusters of severe respiratory illness with clinical presentations greatly resembling SARS-CoV, leading to ICU admission and high mortality^7^. Clinical manifestations have included fever, fatigue, dry cough, shortness of breath, and acute respiratory distress syndrome (ARDS). Additionally, a study of the first 41 laboratory-confirmed cases with COVID-19 showed that 63% of patients had lymphopenia, and cytokine storm could be associated with disease severity. However, little is known about immunologic features between severe and moderate forms of COVID-19^7^.

In this study, we performed a comprehensive evaluation of characteristics of 21 patients with COVID-19 admitted to Tongji Hospital, Wuhan. We aimed to compare the clinical and immunologic features between severe cases and moderate cases. These findings will help us extend our understanding of the pathophysiological mechanism of the SARS-CoV-2 infection.

## Patients and methods

### Study participants

From late Dec 19, 2019 to Jan 27, 2020, a total of 21 cases who initially presented with fever or respiratory symptoms, with pulmonary infiltrates on chest computed tomography (CT) scans in isolation ward of Department of Infectious Disease, Tongji hospital were later confirmed to be infected with the novel coronavirus by the local health authority. Four cases had a history of exposure to the Huanan seafood market.

We retrospectively evaluated and analyzed the medical history, physical examination, and hematological, biochemical, radiological, microbiological and immunological evaluation results obtained from these 21 patients with COVID-19. Epidemiological, clinical, laboratory, and radiological characteristics and treatment as well as outcomes data were obtained from electronic medical records. The data collection forms were reviewed independently by two researchers.

The patients with SpO2≤93% or respiratory rates ≥30 per min were classified as having severe COVID-19, those without above-mentioned signs as moderate cases.

Considering the emergence of COVID-19 cases during the influenza season, oseltamivir (orally 75 mg twice daily) and antibiotics (oral and intravenous) were empirically administered. Corticosteroid therapy (methylprednisolone) was given concomitantly to some patients with COVID-19 by physicians. Oxygen support including nasal cannula and non-invasive mechanical ventilation was provided to the patients according to the severity of hypoxemia. After being identified as having laboratory-confirmed SARS-CoV-2 infection, these patients were transferred to the designated hospital.

The study was approved by the Institutional Review Board of Tongji Hospital, Tongji Medical College, Huazhong University of Science and Technology.

### Complication definitions

ARDS and shock were defined according to the interim guidance of WHO for novel coronavirus^8^.

Hypoxemia was defined as arterial oxygen tension (PaO2) over inspiratory oxygen fraction (FIO2) of less than 300 mm Hg.

Acute kidney injury was identified and classified on the basis of the highest serum creatinine level or urine output criteria according to the kidney disease improving global outcomes classification.

Acute liver injury was defined as jaundice with a total bilirubin level of ≥ 3 mg/dl and an acute increase in alanine aminotransferase of at least five times the upper limit of the normal range and/or an increase in alkaline phosphatase of at least twice the upper limit of the normal range.

Cardiac injury was diagnosed if serum levels of cardiac biomarkers were > the 99th percentile upper reference limit, or new abnormalities were shown in electrocardiography and echocardiography.

Secondary infection including bacteria and fungus was diagnosed if the patients had clinical symptoms or signs of nosocomial pneumonia or bacteremia, and was combined with a positive culture of a new pathogen from a respiratory tract specimen or from blood samples taken ≥48 h after admission.

### Laboratory measurements

#### Respiratory pathogen detection

Respiratory specimens were collected by local CDC and then shipped to designated authoritative laboratories to detect the SARS-CoV-2. The presence of SARS-CoV-2 in respiratory specimens was detected by next-generation sequencing or real-time RT-PCR methods. The primers and probe target to envelope gene of CoV were used and the sequences were as follows: forward primer 5′-TCAGAATGCCAATCTCCCCAAC-3′; reverse primer 5′-AAAGGTCCACCCGATACATTGA-3′; and the probe 5′CY5-CTAGTTACACTAGCCATCCTTACTGC-3′BHQ1. Conditions for the amplifications were 50°C for 15 min, 95°C for 3 min, followed by 45 cycles of 95°C for 15 s and 60°C for 30 s.

#### Clinical laboratory measurements

Initial clinical laboratory investigation included a complete blood count, serum biochemical test (including liver and renal function, creatine kinase, lactate dehydrogenase, and electrolytes), coagulation profile, as well as immunological test (including serum cytokines, peripheral immune cells subsets and the expression of IFN-γ by immune cells). Respiratory specimens, including nasal and pharyngeal swabs, or sputum were tested to exclude evidence of other virus infection, including influenza, respiratory syncytial virus, avian influenza, parainfluenza virus and adenovirus using real-time RT-PCR assays approved by the China Food and Drug Administration. Routine bacterial and fungal examinations were also performed.

#### Cytokine measurement

To explore the impact of SARS-CoV-2 on the secretion of cytokines in the early phase of the infection, plasma cytokines including IL-1β, IL-2R, IL-6, IL-8 (also known as CXCL8), IL-10, and TNF-α were measured using the sandwich enzyme-linked immune-sorbent assay (ELISA) method by micro-ELISA autoanalyser (Diasorin Etimax 3000, Germany) for all patients according to the manufacturer’s instructions. The first initial blood samples were drawn shortly after hospital admission.

#### Proportions analysis of peripheral blood immunological indicators

The proportions and numbers of NK, CD4^+^T, CD8^+^T, Treg and B cells, and the expression of cell surface markers as well as IFN-γ expression by CD4^+^T, CD8^+^T and NK cells were studied in these patients with laboratory-confirmed SARS-CoV-2 infection who were reported by the local health authority. PE-CD28/PE•CY7-CD8/PerCP-CD45/APC-HLADR/APC•CY7-CD3/V450-CD4, FITC-CD45RA/PE-CD45RO/PE•CY7-CD127/PerCP-CD45/APC-CD25/APC•CY7-CD3/V4 50-CD4, FITC-CD3/PE-CD8/PE•CY7-CD56/PerCP-CD45/APC-IFN-γ/APC•CY7-CD3/V450-CD4 multiple-color anti-human monoclonal antibodies (mAbs) combination reagents and matched isotype controls were used to determine the peripheral immune cell subsets within 2 days after admission. Peripheral blood mononuclear cells (PBMCs) were isolated immediately from fresh blood and subjected to antibody staining followed by flow cytometry. All reagents were purchased from Becton, Dickinson, and Company (BD, Franklin Lakes, USA). Cell surface antigen staining for flow cytometry was conducted according to the standard procedure of the BD Pharmingen protocol. All samples were detected by BD FACS Canto II Flow Cytometry System and analyzed with the BD FACS Diva Software.

### Statistical analysis

Continuous variables were expressed as mean (S.D.) and compared with the Mann-Whitney U test; categorical variables were expressed as number (%) and compared by χ2 test or Fisher’s exact test between moderate and severe case groups. A two-sided α of less than 0·05 was considered statistically significant. Statistical analyses were done using the SAS software, version 9.4.

## Results

### Patient demographics and baseline characteristics of severe and moderate forms of COVID-19

By Jan 22, 2020, a total of 21 admitted hospital patients with pneumonia were identified as laboratory-confirmed SARS-CoV-2 infection in Wuhan Tongji hospital. Of these patients, only four patients including a familial cluster of three confirmed cases had direct exposure to Huanan seafood market. 11 (52.4%) patients with SpO2≤93% or respiratory rates ≥30 per min who required high-flow nasal cannula or non-invasive mechanical ventilation using the Bilevel Positive Airway Pressure (BiPAP) mode to correct hypoxemia, were classified as having severe COVID-19, whereas 10 (47.6%) patients without above-mentioned signs as moderate cases. 10 (90.9%) of severe cases and 7 (70%) of moderate cases were male (table 1). The mean age of the severe cases (63.9 years) was significantly older than moderate cases (51.4 years). 90.9% of severe cases were above 50 years old, which was more frequent than that of moderate cases (50%). 5 (45.5%) of severe cases and 2 (20%) moderate cases had underlying diseases. Of 7 patients with underlying diseases, four patients had hypertension, two had diabetes, and one had both hypertension and diabetes. The mean time from onset of symptoms to first hospital admission was 7.3 days in severe cases and 5.5 days in moderate cases.

**Table 1.**
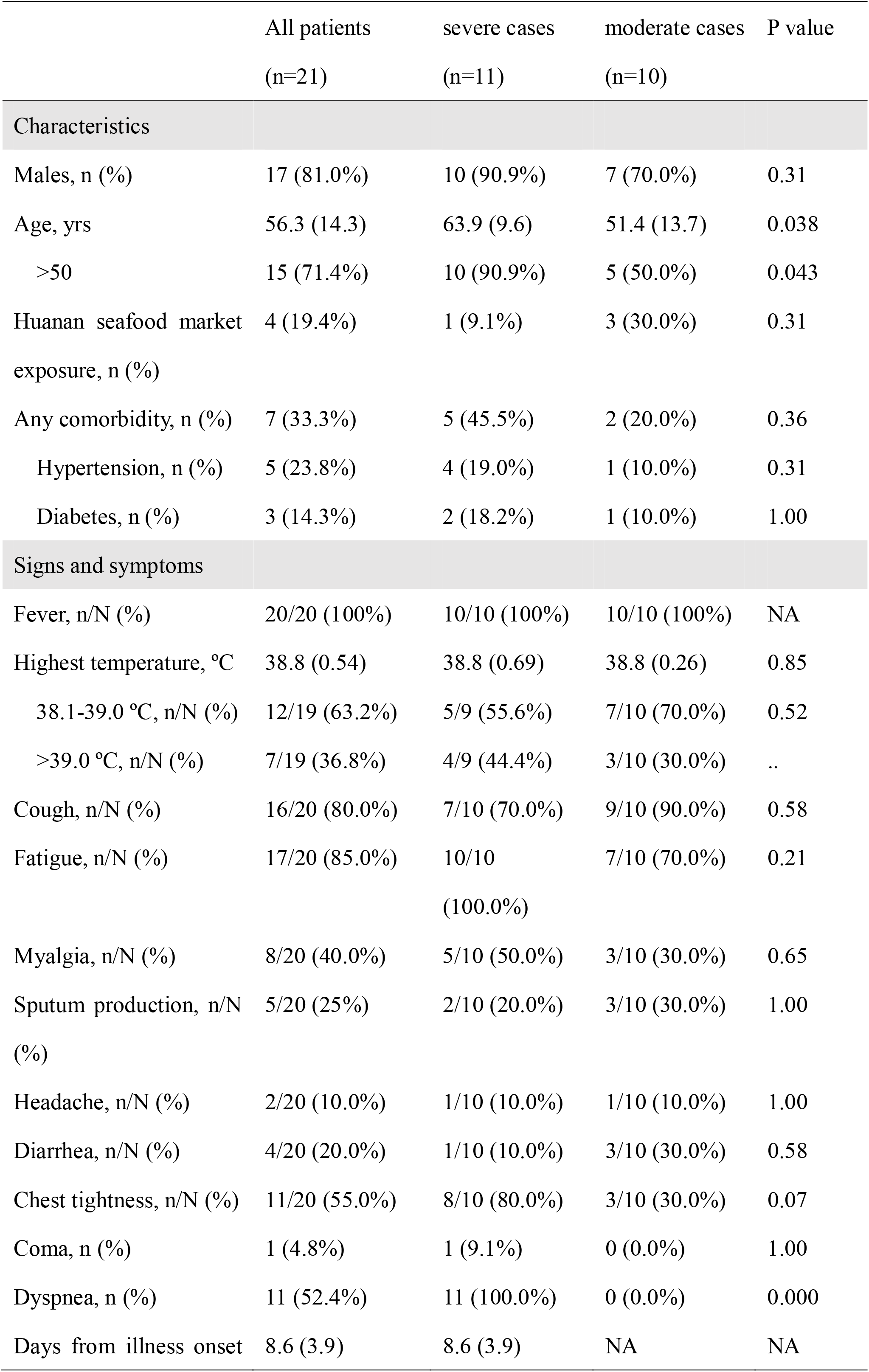
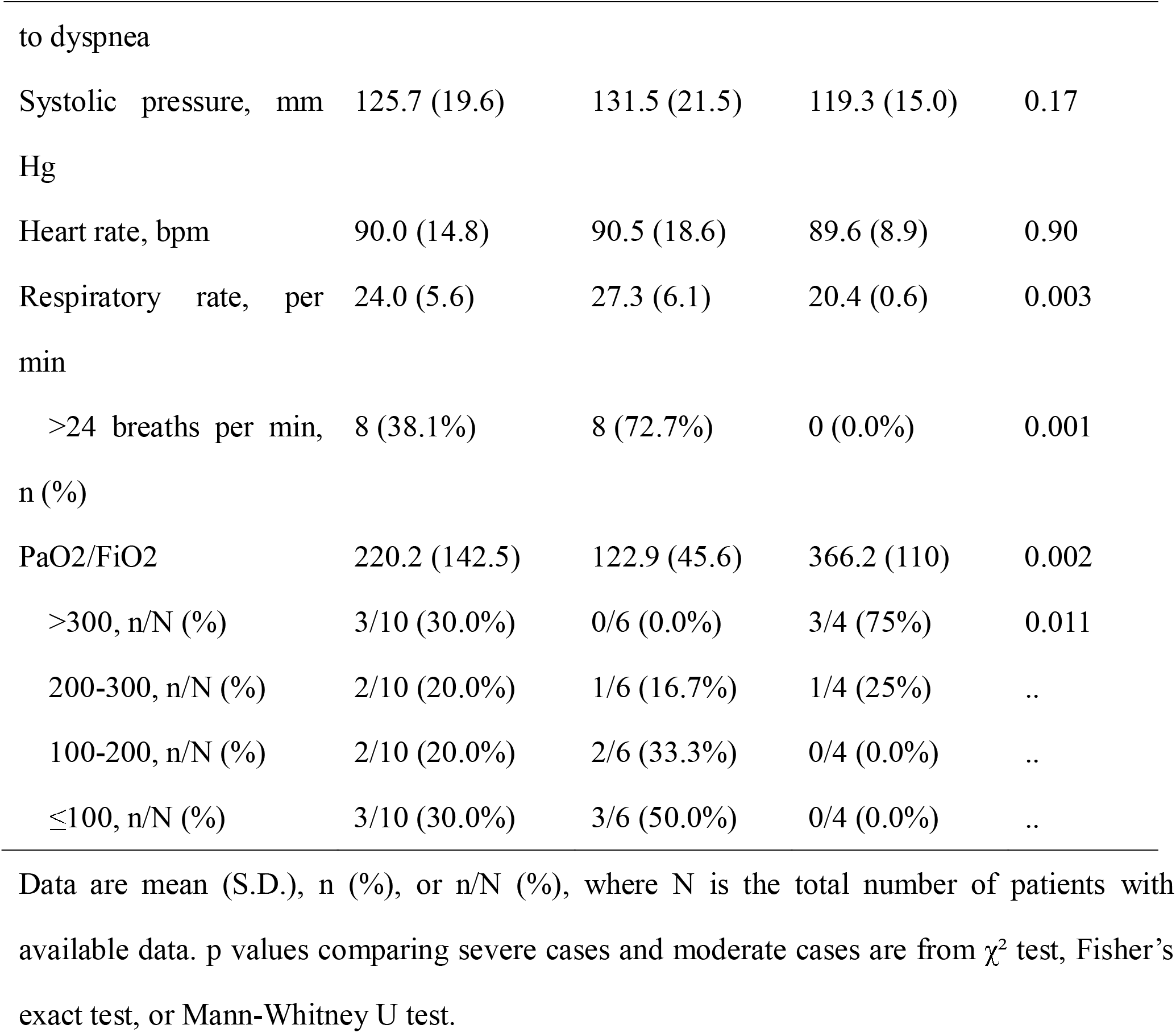
Demographics and baseline characteristics of patients with COVID-19

|  | All patients<br>(n=21) | severe cases<br>(n=11) | moderate cases<br>(n=10) | P value |
| --- | --- | --- | --- | --- |
| **Characteristics** |  |  |  |  |
| Males, n (%) | 17 (81.0%) | 10 (90.9%) | 7 (70.0%) | 0.31 |
| Age, yrs | 56.3 (14.3) | 63.9 (9.6) | 51.4 (13.7) | 0.038 |
| >50 | 15 (71.4%) | 10 (90.9%) | 5 (50.0%) | 0.043 |
| Huanan seafood market exposure, n (%) | 4 (19.4%) | 1 (9.1%) | 3 (30.0%) | 0.31 |
| Any comorbidity, n (%) | 7 (33.3%) | 5 (45.5%) | 2 (20.0%) | 0.36 |
| Hypertension, n (%) | 5 (23.8%) | 4 (19.0%) | 1 (10.0%) | 0.31 |
| Diabetes, n (%) | 3 (14.3%) | 2 (18.2%) | 1 (10.0%) | 1.00 |
| **Signs and symptoms** |  |  |  |  |
| Fever, n/N (%) | 20/20 (100%) | 10/10 (100%) | 10/10 (100%) | NA |
| Highest temperature, °C | 38.8 (0.54) | 38.8 (0.69) | 38.8 (0.26) | 0.85 |
| 38.1-39.0 °C, n/N (%) | 12/19 (63.2%) | 5/9 (55.6%) | 7/10 (70.0%) | 0.52 |
| >39.0 °C, n/N (%) | 7/19 (36.8%) | 4/9 (44.4%) | 3/10 (30.0%) | .. |
| Cough, n/N (%) | 16/20 (80.0%) | 7/10 (70.0%) | 9/10 (90.0%) | 0.58 |
| Fatigue, n/N (%) | 17/20 (85.0%) | 10/10 (100.0%) | 7/10 (70.0%) | 0.21 |
| Myalgia, n/N (%) | 8/20 (40.0%) | 5/10 (50.0%) | 3/10 (30.0%) | 0.65 |
| Sputum production, n/N (%) | 5/20 (25%) | 2/10 (20.0%) | 3/10 (30.0%) | 1.00 |
| Headache, n/N (%) | 2/20 (10.0%) | 1/10 (10.0%) | 1/10 (10.0%) | 1.00 |
| Diarrhea, n/N (%) | 4/20 (20.0%) | 1/10 (10.0%) | 3/10 (30.0%) | 0.58 |
| Chest tightness, n/N (%) | 11/20 (55.0%) | 8/10 (80.0%) | 3/10 (30.0%) | 0.07 |
| Coma, n (%) | 1 (4.8%) | 1 (9.1%) | 0 (0.0%) | 1.00 |
| Dyspnea, n (%) | 11 (52.4%) | 11 (100.0%) | 0 (0.0%) | 0.000 |
| Days from illness onset | 8.6 (3.9) | 8.6 (3.9) | NA | NA |
| to dyspnea |  |  |  |  |
| Systolic pressure, mm Hg | 125.7 (19.6) | 131.5 (21.5) | 119.3 (15.0) | 0.17 |
| Heart rate, bpm | 90.0 (14.8) | 90.5 (18.6) | 89.6 (8.9) | 0.90 |
| Respiratory rate, per min | 24.0 (5.6) | 27.3 (6.1) | 20.4 (0.6) | 0.003 |
| >24 breaths per min, n (%) | 8 (38.1%) | 8 (72.7%) | 0 (0.0%) | 0.001 |
| PaO<sub>2</sub>/FiO<sub>2</sub> | 220.2 (142.5) | 122.9 (45.6) | 366.2 (110) | 0.002 |
| >300, n/N (%) | 3/10 (30.0%) | 0/6 (0.0%) | 3/4 (75%) | 0.011 |
| 200-300, n/N (%) | 2/10 (20.0%) | 1/6 (16.7%) | 1/4 (25%) | .. |
| 100-200, n/N (%) | 2/10 (20.0%) | 2/6 (33.3%) | 0/4 (0.0%) | .. |
| ≤100, n/N (%) | 3/10 (30.0%) | 3/6 (50.0%) | 0/4 (0.0%) | .. |

Data are mean (S.D.), n (%), or n/N (%), where N is the total number of patients with available data. p values comparing severe cases and moderate cases are from $\chi^2$ test, Fisher's exact test, or Mann-Whitney U test.

Four of eleven severe cases died at an average of 20 days after the onset of the illness. Of these four fatal cases, all of them were male and had age over 50 years old, and two had hypertension. Mean age of fatal cases were 67.5 years old. Three of the fatal cases had PaO2/FiO2 ratio ≤ 100 on admission.

Excluding one comatose patient without a clear history (classified as severe case), the most common clinical manifestations at onset of illness include fever (100% of total cases), cough (70% of severe cases, 90% of moderate cases), fatigue (100% of severe cases, 70% of moderate cases) and myalgia (50% of severe cases, 30% of moderate cases). Less common symptoms include sputum production (20% of severe cases, 30% of moderate cases), diarrhea (10% of severe cases, 30% of moderate cases), headache (10% of severe cases, 10% of moderate cases) and hemoptysis (10% of severe cases). All the severe cases developed dyspnea, and nine of them had SpO2<93% even with high-flow nasal cannula, who were then ventilated using the BiPAP mode to treat hypoxemia. The mean duration from illness onset to dyspnea was 8.6 days. Arterial blood gas (ABG) test was performed in 10 patients on admission (six severe and four moderate cases). Of them, PaO2/FiO2 ratio was significantly lower in severe cases (122.9 mmHg) than moderate cases (366.2 mmHg). Three severe cases had PaO2/FiO2 ratio ≤ 100, indicating severe ARDS.

### Laboratory findings and CT scans of severe and moderate forms of COVID-19

The routine blood tests on admission of three (30%) moderate cases showed mild leucopenia (white blood cell count (WBC) < 4 × 10□/L). WBC counts were normal or slightly increased in all the severe cases. Both WBC and neutrophil counts were significantly higher in severe cases (9.2 × 10□/L and 8.0 × 10□/L) than moderate cases (4.7 × 10□/L and 3.1 × 10□/L). Whereas lymphocyte counts were significantly lower in severe cases (0.7 × 10□/L) than moderate cases (1.1 × 10□/L). Lymphopenia (lymphocyte count <0.8 × 10□/L) was found in 8 (72.7%) severe cases and only 1 (10.0%) moderate cases (table 2).

**Table 2.**
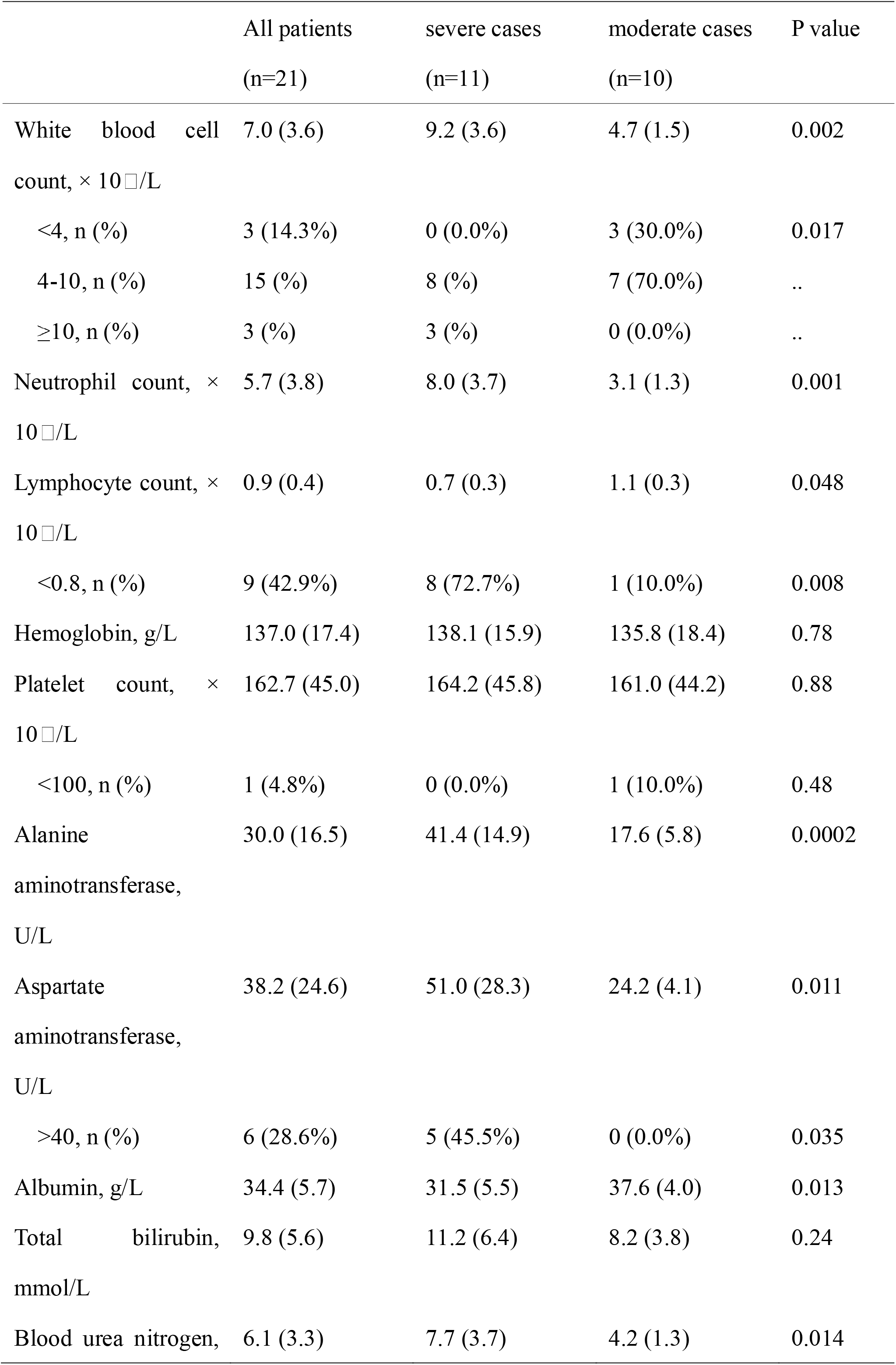
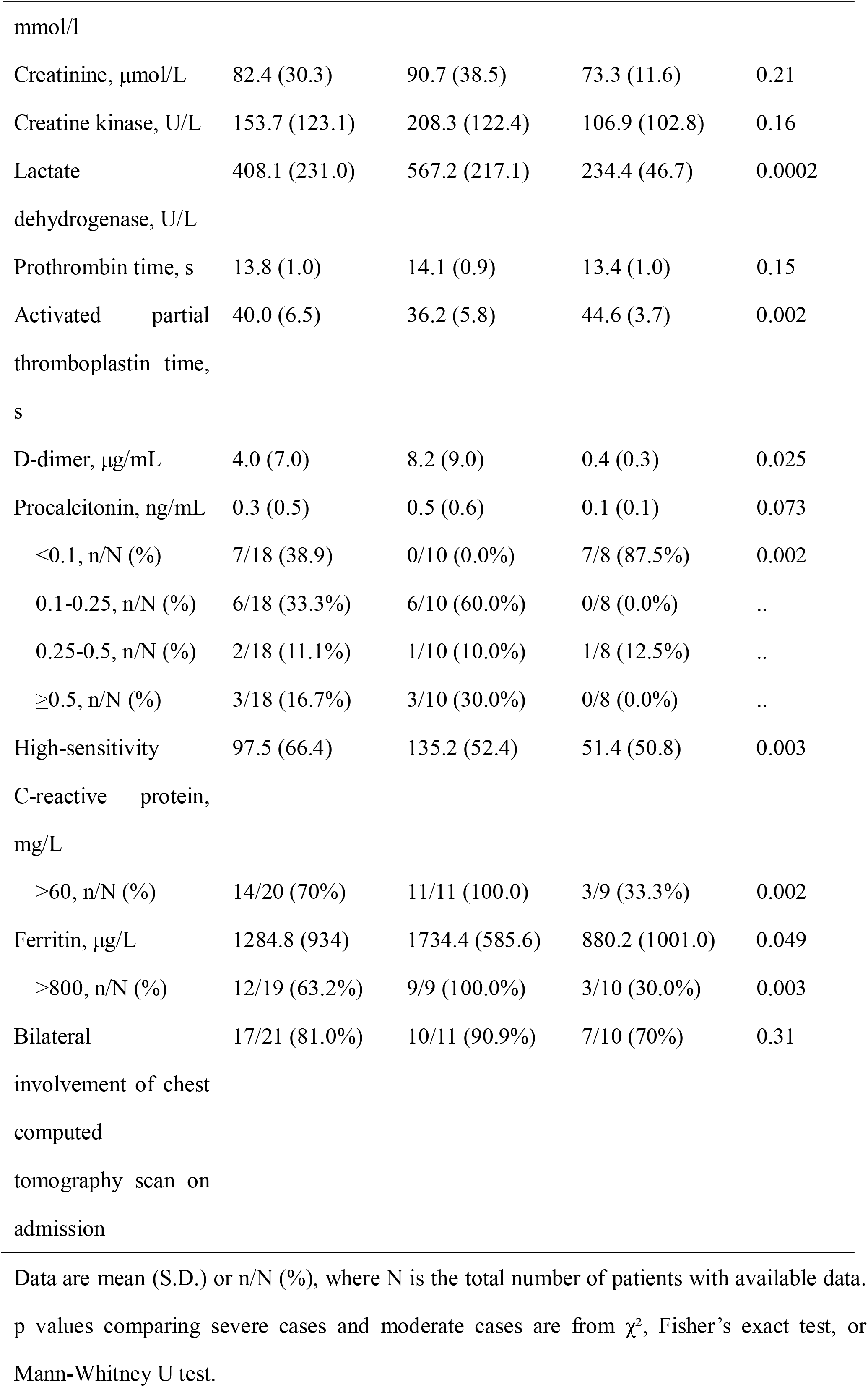
Laboratory findings and chest CT images of patients with COVID-19 on admission

Alanine aminotransferase (ALT) and aspartate aminotransferase (AST) levels were significantly higher in severe cases (41.4 U/L and 51.0 U/L) than moderate cases (17.6 U/L and 24.2 U/L). Albumin concentrations were significantly lower in severe cases (31.5g/L) than moderate cases (37.6g/L). Levels of lactate dehydrogenase (LDH) were significantly higher in severe cases (567.2 U/L) than moderate cases (234.4 U/L).

Concentrations of serum high-sensitivity C-reactive protein (hsCRP) and ferritin on admission were significantly elevated in severe cases (135.2 mg/L and 1734.4 ug/L) than moderate cases (51.4 mg/L and 880.2 ug /L). Serum levels of procalcitonin on admission tended to be higher in severe cases (0.5 ng/mL) than moderate cases (0.1 ng/mL). Activated partial thromboplastin time (APTT) on admission was significantly shorter in severe cases (36.2s) than moderate cases (44.6s). D-dimer levels on admission were markedly greater in severe cases (8.2 ug/mL) than moderate cases (0.4 ug/mL).

Interstitial lung abnormalities were observed in chest CT scans of all patients on admission. Of the 21 patients, 10 (90.1%) severe cases and 7 (70%) moderate cases had bilateral involvement on admission (table 2). Later on, only one patient with moderate COVID-19 remained unilateral involvement. The typical findings of chest CT images of severe COVID-19 on admission showed bilateral ground glass opacity and subsegmental areas of consolidation (figure 1A), then progressed rapidly with mass shadows of high density in both lungs (figure 1B). Whereas the representative chest CT findings of moderate COVID-19 showed bilateral ground glass opacity (figure 1C). Later chest CT images revealed bilateral ground-glass opacity had been resolved (figure 1D).

**Figure 1:**
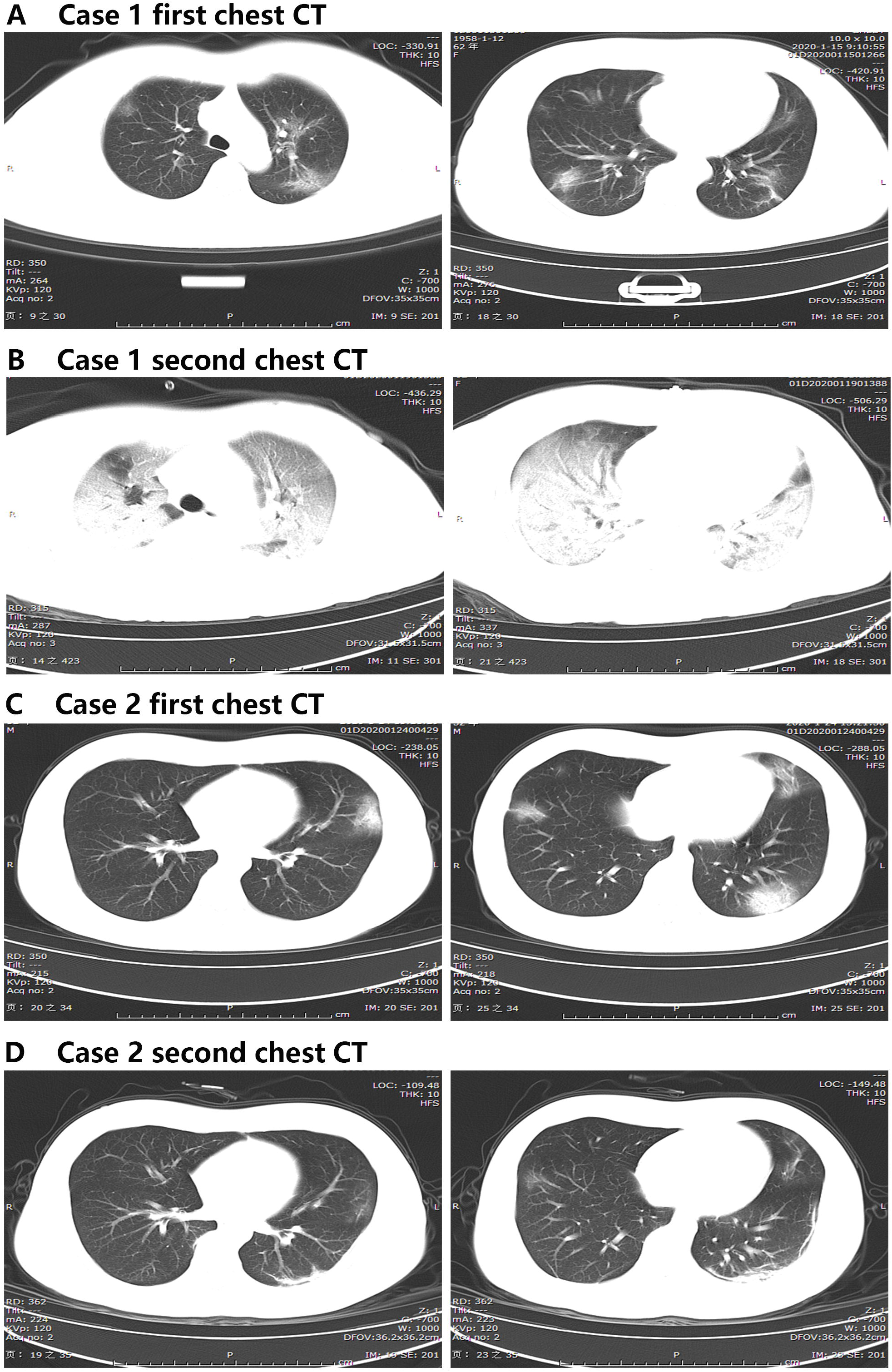
Computed tomography of the chest of patients with COVID-19. Chest CT axial view lung window from a 62-year-old female with severe COVID-19 showing bilateral ground-glass opacity and subsegmental areas of consolidation on day 6 after symptom onset (A), and typical presentation of a “white lung” appearance with bilateral multiple lobular and subsegmental areas of consolidation on day 8 after symptom onset (B). Chest CT axial view lung window from a 32-year-old male with moderate COVID-19 showing bilateral ground-glass opacity on day 7 after symptom onset (C), and resolved bilateral ground-glass opacity on day 11 after symptom onset (D).

### Immunologic features of severe and moderate forms of COVID-19

Evaluation of plasma cytokines revealed that IL-2R, IL-6, IL-10, and TNF-α concentrations on admission were moderately elevated in the majority of severe cases (88.9%, 88.9%, 77.8%, and 88.9%), but only mildly increased (14.3%, 71.4%, 28.6%, and 42.9%) or remained within the normal range in moderate cases (table 3). IL-8 concentrations were elevated in few patients (22.2% severe cases, 14.3% moderate cases). IL-1β concentrations were undetectable (<5pg/mL) in nearly all the patients with either severe or moderate COVID-19. IL-2R, TNF-α and IL-10 concentrations on admission were significantly higher in severe cases (1202.4 pg/mL, 10.9 pg/mL and 10.9 pg/mL) than moderate cases (441.7 pg/mL, 7.5 pg/mL and 6.6 pg/mL).

**Table 3.**
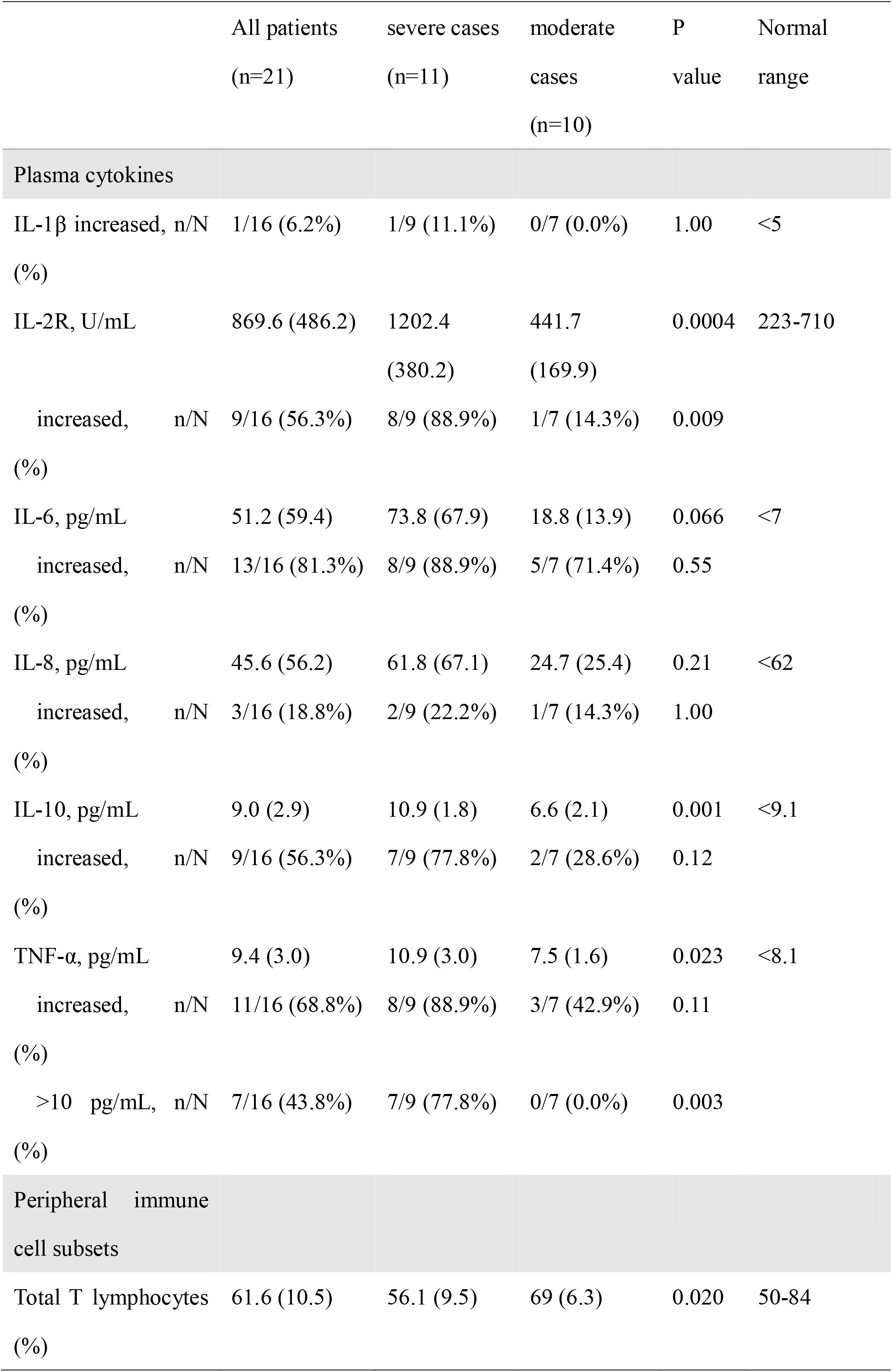
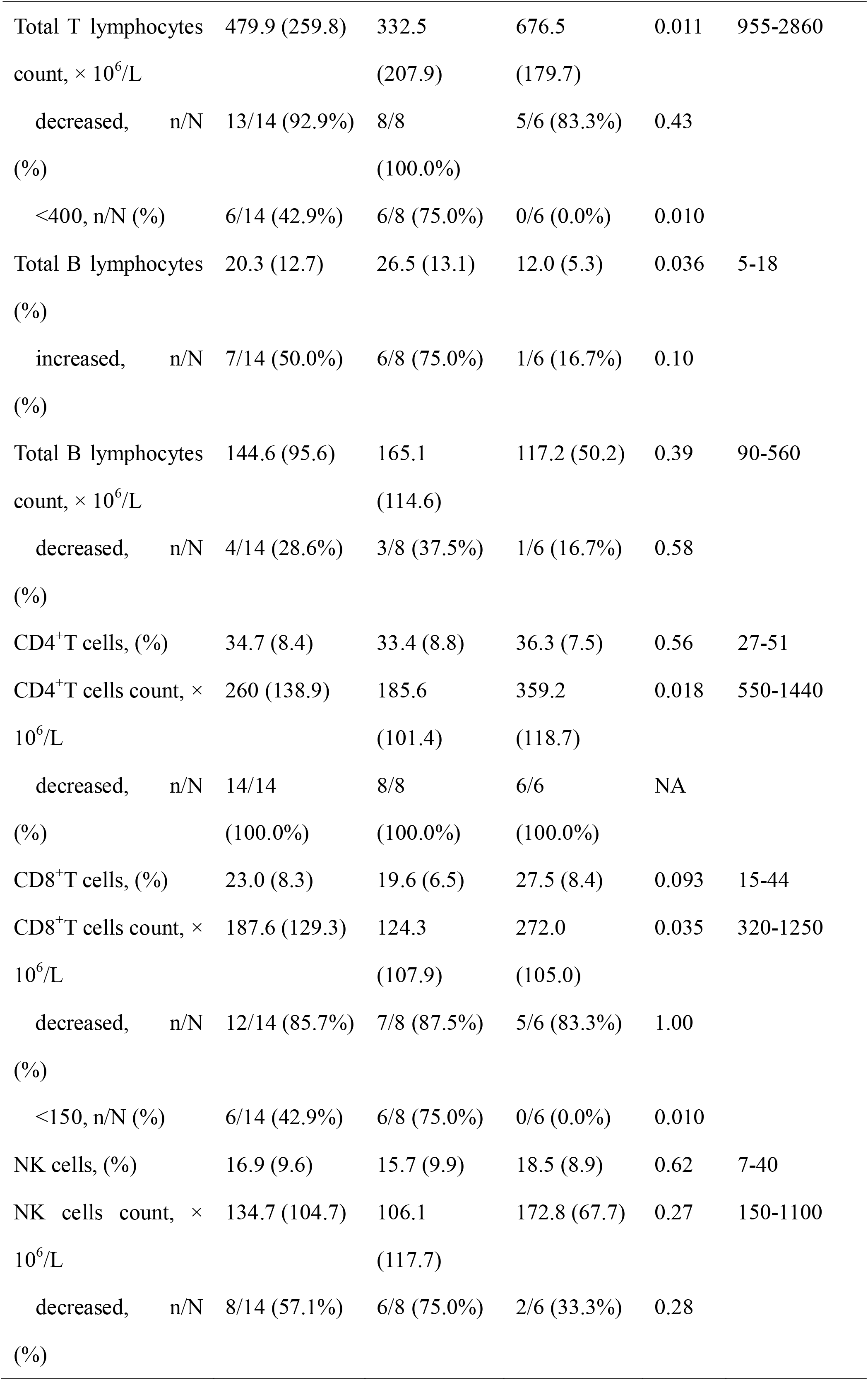
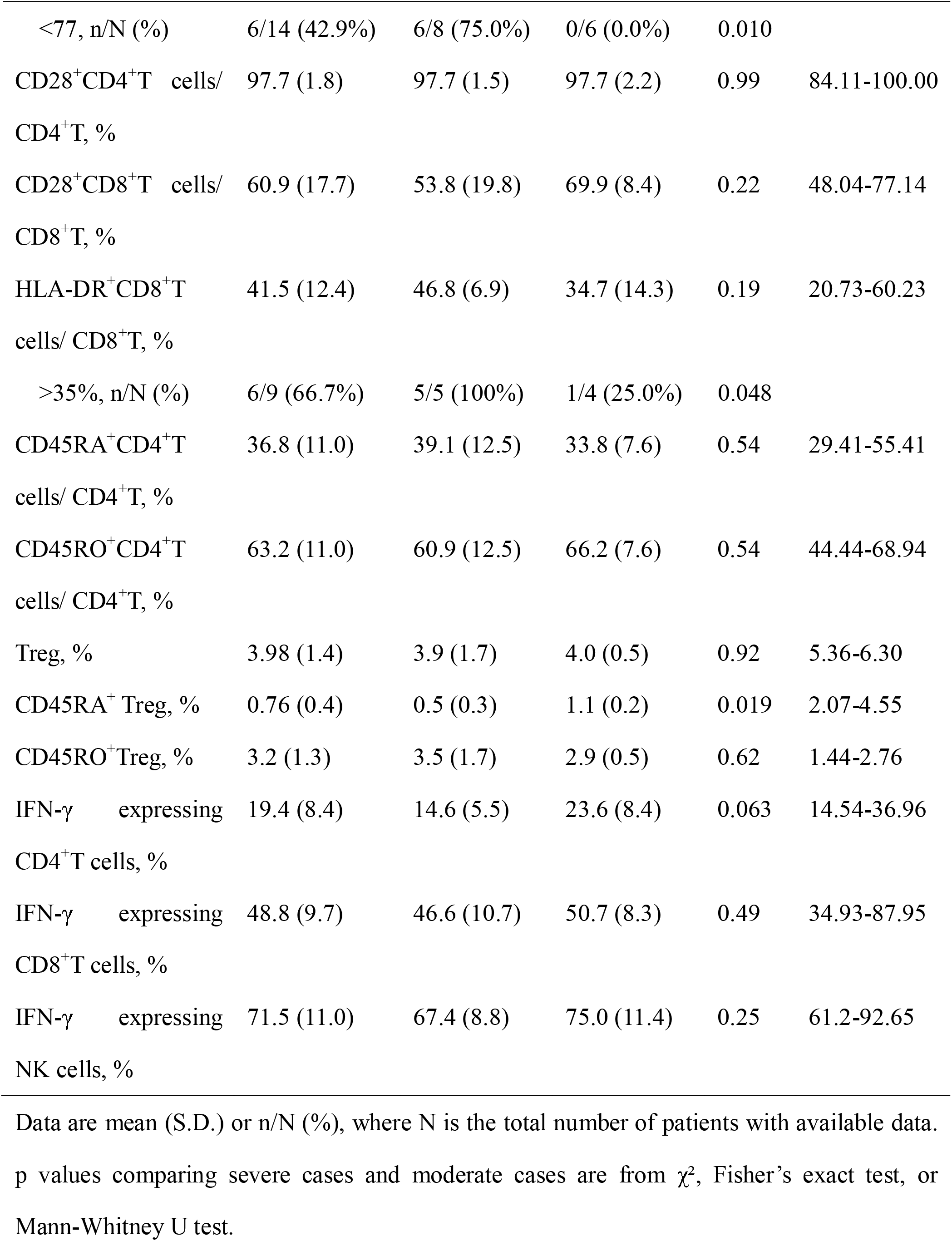
Immunologic features of patients with COVID-19

Preliminary analysis of circulating immune cells subsets showed that absolute number of total T lymphocytes, CD4^+^T cells and CD8^+^T cells decreased in the majority of patients with either severe (100%, 100% and 87.5%) or moderate COVID-19 (83.3%, 100% and 83.3%), and total T lymphocytes, CD4^+^T cells and CD8^+^T cells counts were reduced more profoundly in severe cases (332.5, 185.6 and 124.3 × 10^6^/L) than moderate cases (676.5, 359.2 and 272.0 × 10^6^/L) (figure 2A, 2B). More severe cases (100%) had the proportion of HLA-DR^+^ expression on CD8^+^T cells (of > 35%) than moderate cases (25%). Reduction of B cells and NK cells counts tended to be more frequent in severe cases (37.5% and 75%) than moderate cases (16.7% and 33.3%), whereas the proportion of B cells was significantly higher in severe cases (26.5%) than moderate cases (12.0%). This could be partly due to the more significant decrease of T lymphocytes in severe cases. In addition, six (75.0%) of eight severe cases showed a broad, significant decrease in all the lymphocytes subsets excluding B cells, with total T lymphocytes counts below 400 × 10^6^/L, CD8^+^T cells counts below 150 × 10^6^/L, and NK cells counts below 77 × 10^6^/L. Of these six patients, three (50%) eventually died.

**Figure 2:**
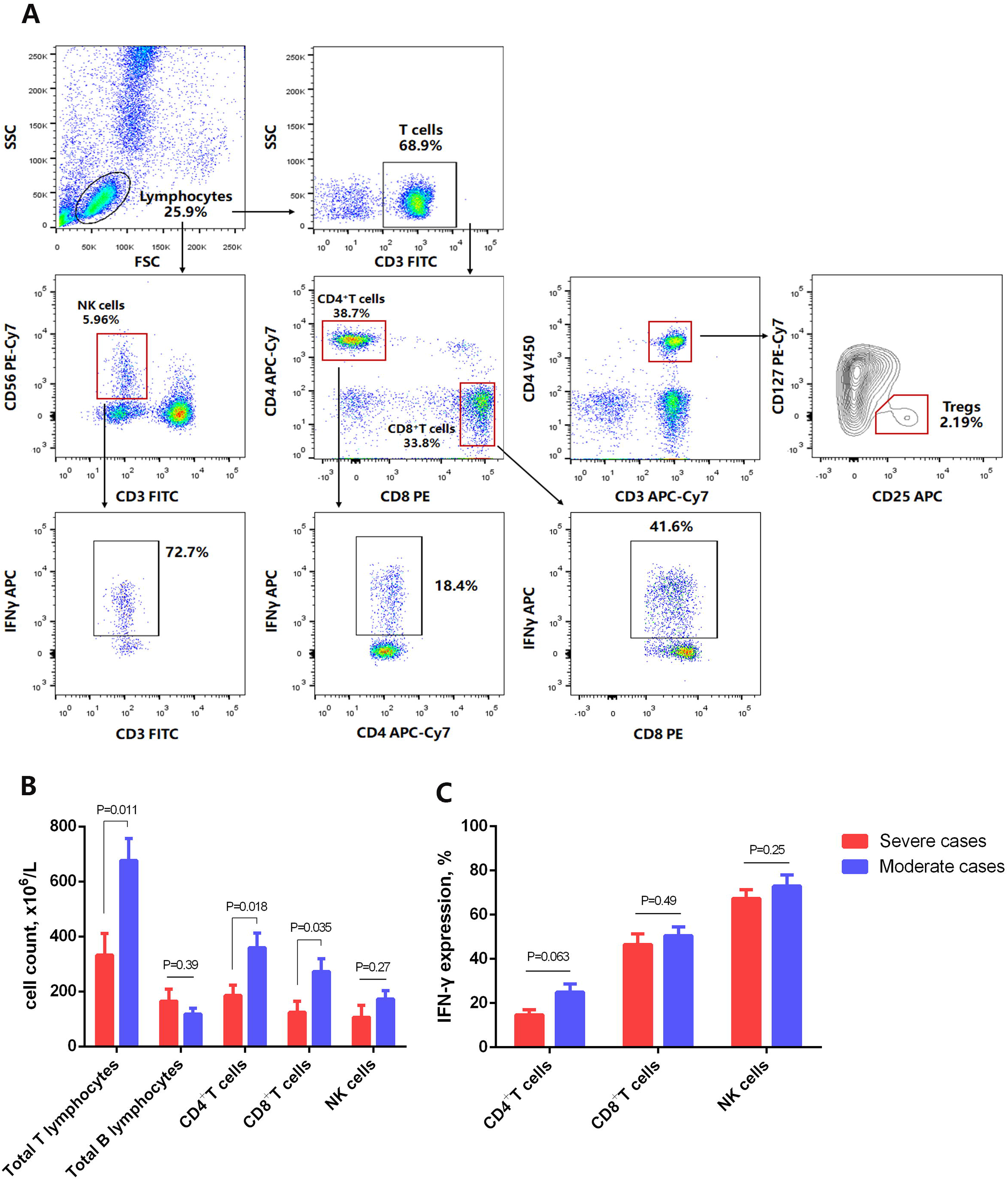
Number of immune cell subsets and proportion of IFN-γ expression in patients with COVID-19. (A) Flow cytometry staining of natural killer (NK) cells, CD4^+^T cells, CD8^+^T cells and Treg as well as production of IFN-γ by CD4^+^T cells, CD8^+^T cells and NK cells from a representative patient. (B) A series of comparisons of absolute number of total T&B lymphocytes, CD4^+^T cells, CD8^+^T cells and NK cells between severe cases and moderate cases. Data are expressed as mean ± SEM. (C) A series of comparisons of production of IFN-γ by CD4^+^T cells, CD8^+^T cells and NK cells between severe cases and moderate cases. Data are expressed as mean ± SEM.

Moreover, the frequencies of Treg (CD4^+^CD25^+^CD127^low+^) and CD45RA^+^Treg decreased in nearly all the severe (60% and 100%) and moderate cases (100% and 100%), with CD45RA^+^Treg decreased more profoundly in severe cases (0.5%) than moderate cases (1.1%). The expressions of IFN-γ by CD4^+^T, CD8^+^T and NK cells were decreased in some patients with severe (50%, 16.7% and 16.7%) or moderate COVID-19 (14.3%, 0% and 14.3%). The expressions of IFN-γ by CD4^+^T cells tended to be lower in severe cases (14.6%) than moderate cases (23.6%) (figure 2C).

### Complications and clinical outcomes of COVID-19

As for complications, nine (81.8%) severe cases and one (10%) moderate case developed ARDS, among them, three (27.3%) severe cases had confirmed severe ARDS by ABG test with PaO2/FiO2 ≤ 100. One case developed multiple organ dysfunction syndrome (MODS), including acute kidney injury, cardiac injury and heart failure as well as pulmonary encephalopathy prior to admission. Another patient developed acute liver injury and disseminated intravascular coagulation in a short time. These two patients died soon afterwards. One patient with severe COVID-19 developed acute liver injury and was in recovery and discharged after a three-week hospitalization, with extracorporeal membrane oxygenation (ECMO) support. The chest CT scan afterwards showed improvement of bilateral ground-glass opacity and subsegmental areas of consolidation.

Of 21 patients, nine (42.9%) required non-invasive ventilation. 19 (90.5%) patients received antiviral therapy (oseltamivir and ganciclovir). All patients were given empirical antimicrobial treatment (moxifloxacin or cefoperazone-sulbactam). In Addition, most patients (85.7%) were administered corticosteroids (methylprednisolone). As of Feb 2, 2020, 4 (36.4 %) of 11 severe cases and none (0.0 %) of the moderate cases have died, the average days from illness onset to death was 20 days. One severe and one moderate case have recovered. Patients were transferred to the designated hospital after being identified as having laboratory-confirmed SARS-CoV-2 infection.

## Discussion

This is the first preliminary study evaluating descriptively the immunologic characteristics of patients with laboratory-confirmed SARS-CoV-2 infection. Both clinical and epidemiological features of patients with COVID-19 have recently been reported^5-7,9^. However, there is insufficient knowledge of pathophysiological parameters as well as immunologic indicators to understand the mechanism involved in COVID-19. Consistent with previous reports^7^, in this present study, a male predominance in the incidence of COVID-19 has been noted similar to that of SARS-CoV, indicating males are more susceptible to SARS-CoV-2 infection than females. Older males (>50 years old), in particular those with chronic comorbidities may be more likely to develop severe COVID-19, and associated with a high mortality (28.6% of males aged >50 years old with comorbidities). The most common clinical manifestations at onset of illness included fever, cough, fatigue and myalgia. Severe cases were more likely to have dyspnea and significantly lower PaO2/FiO2, and to develop ARDS. In terms of laboratory finding, leukocytosis (≥10× 10□/L) but lymphopenia (<0.8× 10□/L) were more common in severe cases than moderate cases. ALT, LDH, D-dimer and inflammatory markers including hsCRP and ferritin were significantly higher in severe cases than moderate cases. Plasma concentrations of both pro-inflammatory cytokines and anti-inflammatory cytokines, including IL-2R, TNF-α and IL-10 increased in the majority of severe cases and was significantly higher than did those in moderate cases, suggesting cytokine storms might be associated with disease severity.

Additionally, we also noted that SARS-CoV-2 infection can cause a significant reduction in peripheral blood lymphocytes and T cell subsets. Although the proportions of T cells subsets in peripheral blood remained within the normal range in most patients, the prevalence of decreasing CD4^+^T cell counts and CD8^+^T cell counts was considerably high in both severe and moderate cases. It is notable that both the proportion and number of B cells were not frequently decreased in these patients. More importantly, the number of CD4^+^T cells and CD8^+^T cells was markedly lower in severe cases than moderate cases. Albeit diminished CD8^+^ T cells, the proportion of HLA-DR^+^ expression on CD8^+^T cells over 35% was more common in severe cases than those of moderate cases. The expression of HLA-DR on human T cells has been regarded primarily as a marker of activated T cells. However, It has been recently indicated that CD8^+^ HLA-DR^+^ T cells constitute to a natural subset of Treg and may exert suppressive effect involving CTLA-4 signaling between neighboring T cells ^10^. Besides, six out of eight severe cases and none of moderate cases with available immunologic data exhibited a broad, significant decline in all the lymphocyte subsets excluding B cells, with total T lymphocyte counts below 400 × 10^6^/L, CD8^+^T cells below 150 × 10^6^/L, and NK cells below<77 × 10^6^/L, of these six patients, three (50%) eventually died, indicating that SARS-CoV-2 infected patients with severe lymphopenia have high-risk of developing severe COVID-19 and extremely high mortality. Moreover, the production of IFN-γ by CD4^+^T cells but not CD8^+^T cells or NK cells tended to be lower in severe cases than moderate cases. These data suggest that SARS-CoV-2 infection may affect primarily T lymphocytes, particularly CD4^+^T cells, resulting in diminished number as well as their IFN-γ production, which might be correlated with disease severity of COVID-19.

CD4^+^ T cells play a pivotal role in regulating immune responses, orchestrating the deletion and amplification of immune cells, especially CD8^+^ T cells. CD4^+^ T cells facilitate virus-specific antibody production via the T-dependent activation of B cells^11^. However, CD8^+^ T cells exert their effects mainly through two mechanisms, cytolytic activities against target cells or cytokines secretion, including IFN-γ, TNF-α, and IL-2 as well as many chemokines^12^.

The production of IFN-γ is essential for the resistance against infection of various pathogens such as virus, bacteria, and parasite^13^. As a major source of IFN-γ, the ability of T cells to respond to infection is part of the adaptive immune response and takes days to develop a prominent IFN-γ response.

The roles of T cell responses in the context of SARS-CoV and MERS-CoV infection have been previously studied. Likewise, patients who survived SARS-CoV and MERS-CoV infections usually had better immune responses than those who did not^14^. The immune system plays an important role in both diseases, but it is differentially affected by the two viruses^15^. A study in SARS-CoV mice model has shown that depletion of CD8^+^ T cells at the time of infection does not affect viral replication or clearance. However, depletion of CD4^+^ T cells leads to an enhanced immune-mediated interstitial pneumonitis and delayed clearance of SARS-CoV from the lungs, demonstrating the vital role of CD4^+^ T but not CD8^+^ T cells in primary SARS-CoV infection ^16^. A Chinese study in SARS-CoV-infected patients has demonstrated that the majority of infiltrative inflammatory cells in the pulmonary interstitium are CD8^+^ T cells that play an important role in virus clearance as well as in immune-mediated injury^17^. After comparing T cell-deficient mice and B cell-deficient mice, it is found that T cells are able to survive and kill virus-infected cells in the MERS-CoV infected lung ^18^. These data highlight the importance of T lymphocytes, in particular, CD4^+^ T cells but not B cells in controlling and finetuning the pathogenesis and outcomes of SARS-CoV and MERS-CoV infection.

Hin Chu et al demonstrated that MERS-CoV but not SARS-CoV can efficiently infect T cells from the peripheral blood and from human lymphoid organs, and induce apoptosis in T cells, which involves the activation of both the extrinsic and intrinsic apoptosis pathways. This may partly explain the lymphopenia observed in MERS-CoV-infected patients^19^. The inability of the SARS-CoV to infect T cells may be ascribed to the lower expression of the SARS-CoV receptor, angiotensin-converting enzyme 2 expression (ACE2) in T cells^15,19^. Several recent structural analyses predict that SARS-CoV-2 also uses ACE2 as its host receptor ^20^. Therefore, these findings indicate that T cells might not be susceptible to SARS-CoV-2 infection and warrants further investigation. Nevertheless, similar to SARS-CoV-2 infection, SARS-CoV can also significantly decrease peripheral CD4^+^ and CD8^+^ T lymphocyte subsets and it was related to the onset of illness ^21^. Several potential mechanisms may be involved, including the development of auto-immune antibodies or immune complexes triggered by viral infection, directly infecting and promoting the growth inhibition and apoptosis of hematopoietic stem and progenitor cells, as well as the use of glucocorticoids, which may account for the decrease of lymphocytes in some SARS patients^22^.

Although nothing is known about mechanism underlying the lymphopenia caused by SARS-CoV-2 infection, in this present study, some patients were administered methylprednisolone, the average dosage was a bit higher in severe cases (40-80mg once a day) than moderate cases (20-40mg once a day). Since corticosteroids have a profound effect on circulating T lymphocytes, which may involve their movement out of the intra-vascular compartment^23^. In this study, we could not exclude the possibility that some of the lymphopenia may be associated with use of steroids, but it does not account for all the cases, and certainly not for the lymphopenia at the initial presentation. Further research is required to determine the effects of corticosteroid on lymphocytes in the patient with COVID-19.

Our study has some limitations. First of all, we mainly evaluated the number of T cell subsets and NK cells as well as their IFN-γ production, the function of these cells, and the role of activated macrophages and lymphocytes infiltrating lung parenchyma remain unclear. Second, this study only included a small number of patients, thus the results should be interpreted with caution, and statistical non-significance may not rule out difference between severe and moderate cases. Third, since data regarding the viremia profile of SARS-CoV-2 are not available, further studies are needed to investigate the correlation between the virus load kinetics and the dynamics of cellular immune responses. Clarification of these questions will allow further dissection of the complex SARS-CoV-2 pathogenesis, with potential implications for the development of therapeutics and vaccines.

In conclusion, the SARS-CoV-2 infection may affect primarily T lymphocytes, especially CD4^+^T cells, resulting in significant decrease in number as well as IFN-γ production, which may be associated with disease severity. Together with clinical characteristics, early immunologic indicators including diminished T lymphocytes and elevated cytokines may serve as potential markers for prognosis in COVID-19. Gaining a deeper understanding of the factors that influence lymphocytes particularly CD4^+^T cell counts and their decrease in patients with SARS-CoV-2 infection is of importance for clinical management of COVID-19.

## Data Availability

All data generated or used during the study are available from the corresponding author by request

## Contributors

QN designed the study and had full access to all data in the study and take responsibility for the integrity of data and the accuracy of the data analysis.

GC and DW contributed to patient recruitment, data collection, data analysis, data interpretation, literature search, and writing of the manuscript.

WG and YC had roles in patient recruitment, data collection, and clinical management.

DH, HW, TW and XZ had roles in the experiments, data collection, data analysis, and data interpretation.

All authors contributed to data acquisition, data analysis, or data interpretation, and reviewed and approved the final version of the manuscript.

## Declaration of interests

All authors declare no competing interests.

## Acknowledgments

This work is funded by grants from Tongji Hospital for Pilot Scheme Project, and partly supported by the Chinese National Thirteenth Five Years Project in Science and Technology (2017ZX10202201).

## Notes

### Competing Interest Statement

The authors have declared no competing interest.

